# Major bleeding increases the risk of subsequent cardiovascular events in patients with atrial fibrillation: Insights from the SAKURA AF registry and RAFFINE registry

**DOI:** 10.1101/2023.06.26.23291925

**Authors:** Hideki Wada, Katsumi Miyauchi, Satoru Suwa, Sakiko Miyazaki, Hidemori Hayashi, Yuji Nishizaki, Naotake Yanagisawa, Katsuaki Yokoyama, Nobuhiro Murata, Yuki Saito, Koichi Nagashima, Naoya Matsumoto, Yasuo Okumura, Tohru Minamino, Hiroyuki Daida

## Abstract

**Background:** Bleeding events are one of the major concerns in patients using oral anticoagulants (OACs). We aimed to evaluate the association between major bleeding and long-term clinical outcomes in atrial fibrillation (AF) patients taking OACs.

**Methods:** We analyzed a database comprising two large-scale prospective registries of patients with documented AF: the RAFFINE and SAKURA registries. The primary outcome was major adverse cardiac and cerebrovascular events (MACCE), defined as the composite of all-cause death, ischemic stroke, and myocardial infarction. Major bleeding was defined in accordance with the criteria of the International Society on Thrombosis and Haemostasis. Cox multivariate analysis was used to determine the impact of major bleeding on the incidence of MACCE.

**Results:** The median follow-up period was 39.7 months. Among 6,633 patients with AF who were taking OAC, 298 (4.5%) had major bleeding and 737 (11.1%) had MACCE. The incidence of MACCE was higher in patients with bleeding than in those without (18.33 and 3.22, respectively, per 100 patient-years; log-rank p <0.0001). Multivariate logistic regression analysis revealed older age, vitamin K antagonist use, and antiplatelet drug use as independent predictors of major bleeding. Multivariate Cox hazard regression analysis showed that the risk of MACCE was significantly higher in patients with major bleeding compared to those without (hazard risk, 4.64; 95% confidence interval, 3.62–5.94; p <0.0001).

**Conclusions:** Major bleeding was associated with long-term adverse cardiovascular events among AF patients taking OAC. Therefore, reducing the risk of bleeding is important for improving clinical outcomes in patients with AF.

## Introduction

Atrial fibrillation (AF) is the most common sustained arrhythmia in clinical practice, and its rate of incidence continues to increase ^1, 2^. Overall, AF increases the risk of thromboembolic stroke; therefore, patients with AF often require treatment with an oral anticoagulant (OAC) such as vitamin K antagonists (VKAs) or direct oral anticoagulants (DOACs) ^2^. Recent guidelines have recommended DOACs over VKAs ^2^. In a recent meta-analysis, DOACs were associated with significant reductions of 19% for the risk of stroke/systemic embolism and 51% for hemorrhagic stroke, compared with VKAs ^3^. However, bleeding events remain a major concern even in patients using DOACs because major bleeding is not uncommon and is associated with adverse cardiovascular events ^4–6^. A sub-analysis of the ARISTOTLE trial demonstrated that major bleeding in patients taking OACs increased the risk of mortality, stroke, or myocardial infarction (MI) within 30 days after major bleeding events ^7^. However, this sub-analysis demonstrated the relationship between bleeding and adverse events only in the short term, and there are few reports regarding whether bleeding is involved in long-term cardiovascular events. The aim of the present study was to evaluate long-term clinical outcomes after a major bleeding event in patients with AF who were taking OACs, using data from two Japanese multicenter, prospective registries.

## Methods

### Study subjects

The analyzed database included two large-scale prospective registries of patients with documented AF: the RAFFINE ^8^ and SAKURA ^9^ registries.

The RAFFINE registry is an observational, multicenter, prospective registry of Japanese patients with AF. It was designed to follow clinical events for at least 3 years and up to 5 years (UMIN Clinical Trials Registry: UMIN000009617) ^8^. In total, 3,901 patients were enrolled between January 2013 and December 2015 at 5 Juntendo University hospitals and 50 general hospitals and clinics. Follow-up was completed in December 2018^10^. Patients aged 20 years or older with a diagnosis of AF by 12-lead electrocardiogram (ECG) or 24-hour Holter ECG were eligible for enrollment into the RAFFINE registry. Physicians at each site enrolled consecutive patients with AF who were under regular clinical observation (for at least 3 months) at the outpatient department. Patients with a life expectancy <1 year, hospital in-patients, and those who failed to provide written informed consent were not enrolled.

The SAKURA is also a prospective observational registry focusing on patients with documented AF being treated with anticoagulants ^9^. Overall, 3,266 patients were enrolled at 2 university hospitals, 13 affiliated or community hospitals, and 48 private clinics between September 2013 and December 2015 (UMIN Clinical Trials Registry: UMIN000014420).

Patients aged ≥20 years with AF were diagnosed using 12-lead ECG, Holter ECG, or event-activated ECG in all patients. Patients with rheumatic mitral valve disease, those with a history of prosthetic valve replacement, those with active infectious endocarditis, and those who failed to provide written informed consent were excluded.

Ethics approval was obtained from the relevant ethics committee for each registry (approval numbers: M11-0799 [11 November 2014] for RAFFINE and RK-130111-2 [1 February 2013] for SAKURA). The Institutional Review Board of our institution approved the study protocol for analysis of the combined dataset of the two cohorts. All patients provided written informed consent. The study was conducted in accordance with the ethical principles of the Declaration of Helsinki.

### Bleedings and endpoints

Major bleeding was defined in accordance with the criteria of the International Society on Thrombosis and Haemostasis (ISTH) as follows: (i) a reduction in hemoglobin level of at least 2 g/dL; (ii) transfusion of at least two units of blood; (iii) symptomatic bleeding at a critical area or organ ^11^.

The primary outcome in this study was major adverse cardiac and cerebrovascular events (MACCE), defined as the composite of all-cause death, ischemic stroke, and myocardial infarction. Secondary outcomes were all-cause death and cardiovascular death. Clinical outcome events were reported by physicians and verified by document submission as much as possible.

### Statistical analysis

Quantitative data are presented as the mean ± standard deviation (SD) or the median (interquartile range [IQR]). Categorical variables are presented as frequencies. Continuous variables were compared using an unpaired t-test or the Mann–Whitney U test. Categorical variables (presented as frequencies) were compared using the chi-squared test or Fisher’s exact probability test. Logistic regression analysis was performed to clarify the factors associated with the incidence of a major bleeding event. Variables that showed an unadjusted value of p <0.05 among potential confounders (age, sex, body mass index, history of major bleeding events, use of VKAs, use of antiplatelet drugs, serum creatinine, hypertension, and diabetes mellitus) were included in the multivariate logistic model.

The Kaplan–Meier method was used to estimate cumulative event rates, and differences in the incidence rates (shown as percentages per patient-year) were analyzed using the log-rank test. A Cox proportional-hazards model was used to compare outcomes between the groups, with results expressed as the hazard ratio (HR) with 95% confidence interval (CI). To investigate the risk of MACCE in patients with bleeding, we compared the incidence of MACCE in patients with and without bleeding during follow-up. Cox multivariate analysis was performed to determine the impact of major bleeding on the incidence of MACCE. The following variables were entered in the multivariate model: age, sex, body mass index, hypertension, diabetes mellitus, use of VKAs, and congestive heart failure. The incidence risk of secondary outcomes were analyzed in the same manner, and variables showing values of p < 0.05 were considered statistically significant. All statistical analyses were performed using JMP version 14.2.0 software (SAS Institute, Cary, NC, USA).

## Results

### Patients and baseline characteristics

After combining the RAFFINE and SAKURA registry data, we enrolled 7,167 patients with AF. Excluded were those without follow-up data (N = 54) and those not taking OAC (N = 480). A final total of 6,633 patients were assessed in the present study **(Figure 1)**. The median follow-up period was 39.7 (IQR, 33.1–48.1) months. Mean patient age was 72 years, 71% were male, 36% had paroxysmal AF, mean CHADS_2_ score was 1.9, and 20% were taking antiplatelet drugs in addition to OACs.

**Figure 1.**
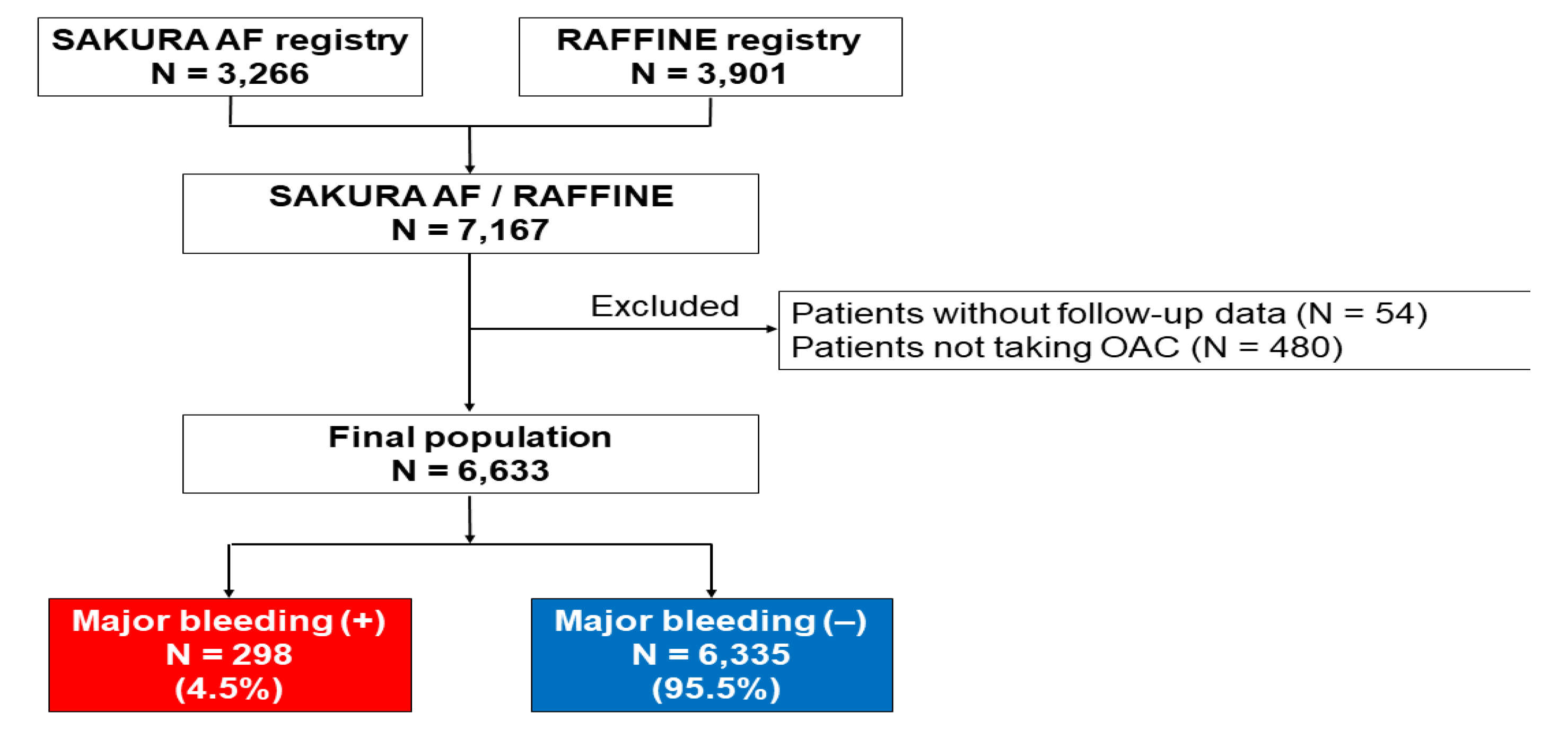
Flow chart of patient selection After the RAFFINE (N = 3,901) and SAKURA (N = 3,266) registry data were combined, 7,167 patients with AF were enrolled. After excluding patients without follow-up data (N = 54) and those who were not taking OAC (N = 480), a final total of 6,633 patients were assessed in the present study. During follow-up, 298 (4.5%) patients had a major bleeding event. AF; atrial fibrillation, OAC; oral anticoagulant.

Among the 6,633 patients, 298 (4.5%) had a major bleeding event during follow-up, with incidence of 1.38 per 100 patient-years. **Table 1** lists the patient characteristics at baseline, stratified by occurrence of a major bleeding event during the follow-up period. Patients with an incident of major bleeding were significantly older, more likely to have ischemic heart disease and previous stroke, and were more often taking VKA and antiplatelet drugs than those without an incident of major bleeding. Baseline CHADS_2_, CHA_2_DS_2_-VASc, and HAS-BLED score were significantly higher and hemoglobin level was significantly lower in patients with major bleeding.

**Table 1.**
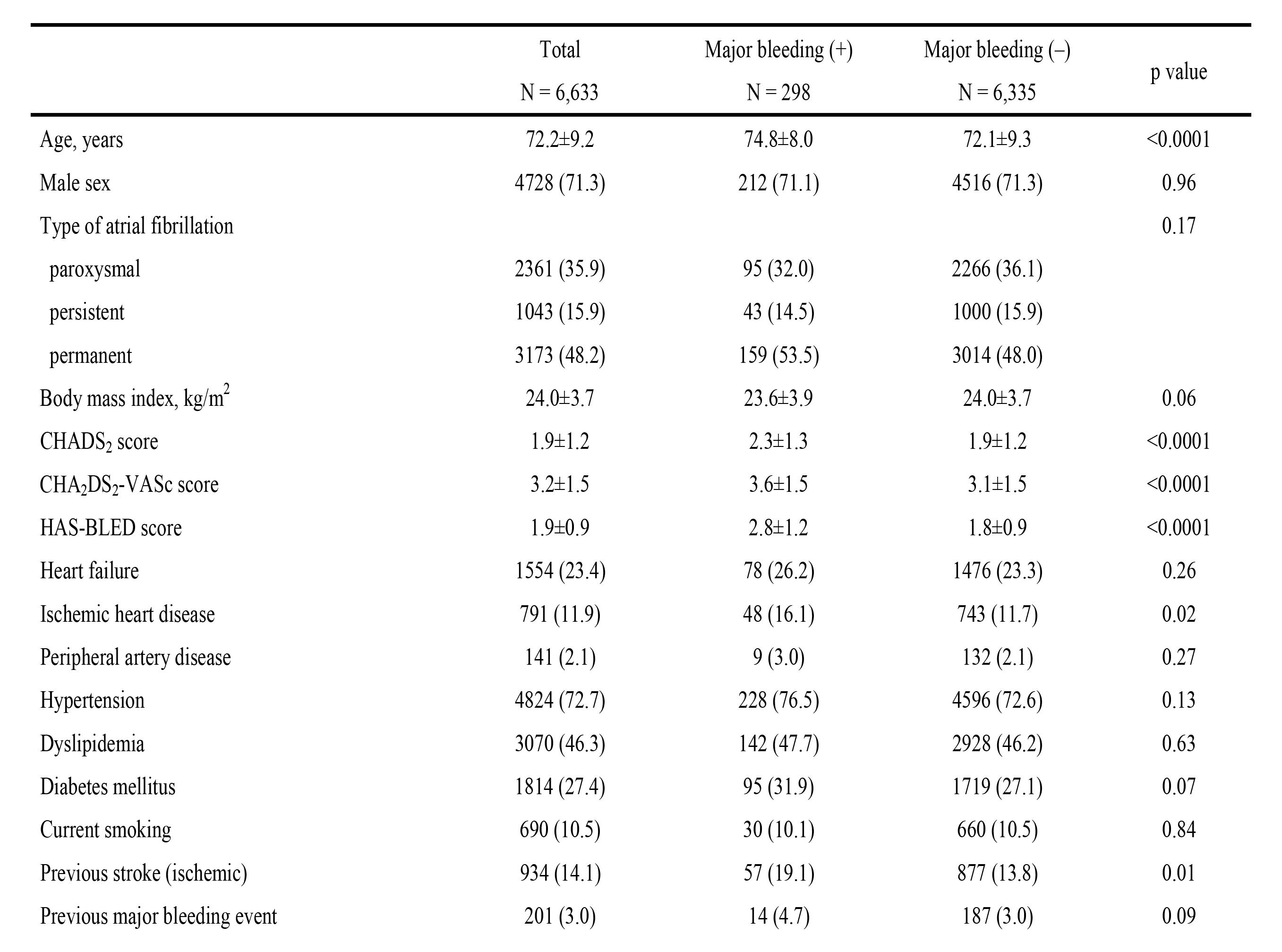
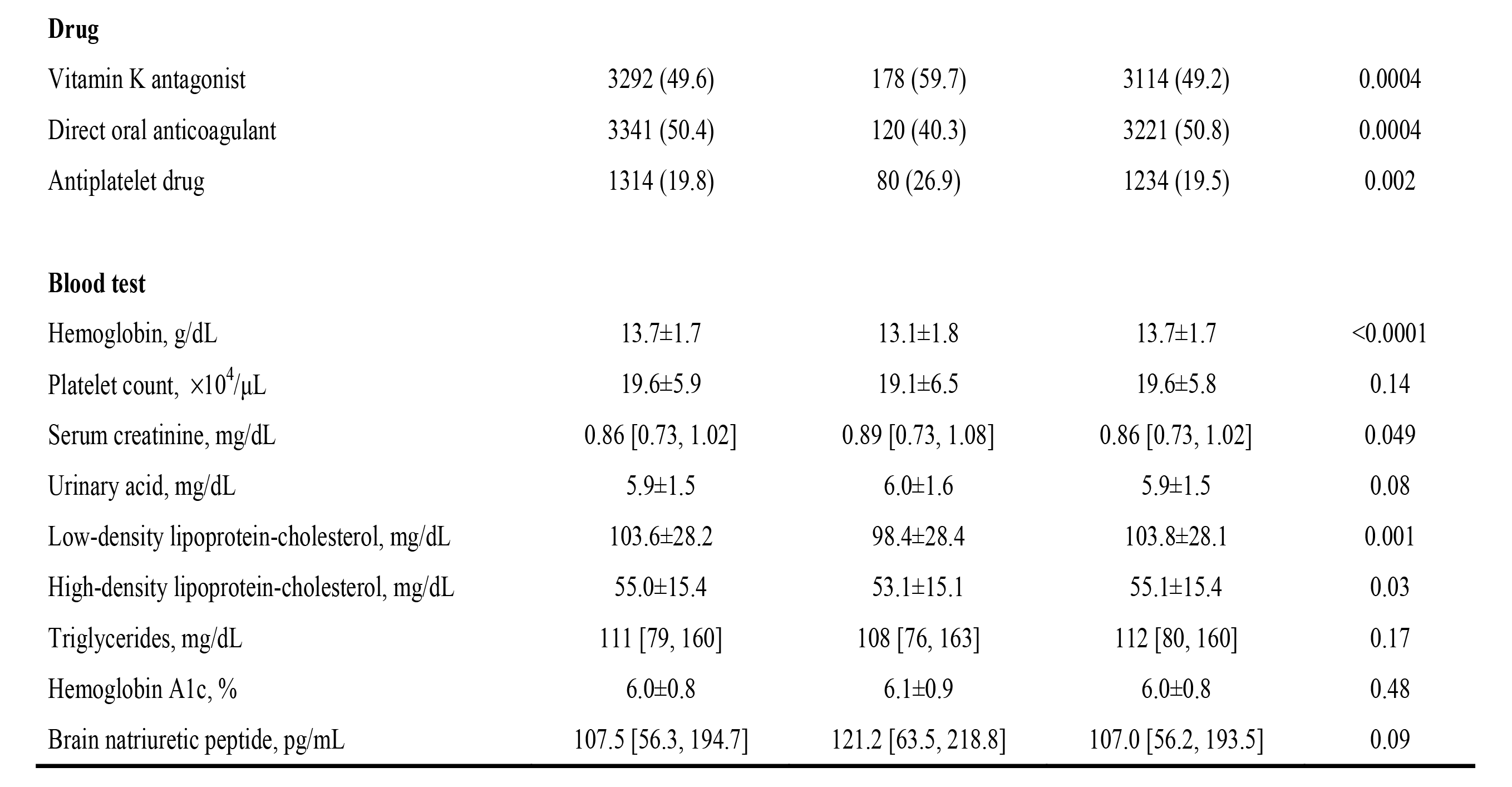
Baseline clinical characteristics

|  | Total<br>N = 6,633 | Major bleeding (+)<br>N = 298 | Major bleeding (-)<br>N = 6,335 | p value |
| --- | --- | --- | --- | --- |
| Age, years | 72.2±9.2 | 74.8±8.0 | 72.1±9.3 | <0.0001 |
| Male sex | 4728 (71.3) | 212 (71.1) | 4516 (71.3) | 0.96 |
| Type of atrial fibrillation |  |  |  | 0.17 |
| paroxysmal | 2361 (35.9) | 95 (32.0) | 2266 (36.1) |  |
| persistent | 1043 (15.9) | 43 (14.5) | 1000 (15.9) |  |
| permanent | 3173 (48.2) | 159 (53.5) | 3014 (48.0) |  |
| Body mass index, kg/m<sup>2</sup> | 24.0±3.7 | 23.6±3.9 | 24.0±3.7 | 0.06 |
| CHADS<sub>2</sub> score | 1.9±1.2 | 2.3±1.3 | 1.9±1.2 | <0.0001 |
| CHA<sub>2</sub>DS<sub>2</sub>-VASc score | 3.2±1.5 | 3.6±1.5 | 3.1±1.5 | <0.0001 |
| HAS-BLED score | 1.9±0.9 | 2.8±1.2 | 1.8±0.9 | <0.0001 |
| Heart failure | 1554 (23.4) | 78 (26.2) | 1476 (23.3) | 0.26 |
| Ischemic heart disease | 791 (11.9) | 48 (16.1) | 743 (11.7) | 0.02 |
| Peripheral artery disease | 141 (2.1) | 9 (3.0) | 132 (2.1) | 0.27 |
| Hypertension | 4824 (72.7) | 228 (76.5) | 4596 (72.6) | 0.13 |
| Dyslipidemia | 3070 (46.3) | 142 (47.7) | 2928 (46.2) | 0.63 |
| Diabetes mellitus | 1814 (27.4) | 95 (31.9) | 1719 (27.1) | 0.07 |
| Current smoking | 690 (10.5) | 30 (10.1) | 660 (10.5) | 0.84 |
| Previous stroke (ischemic) | 934 (14.1) | 57 (19.1) | 877 (13.8) | 0.01 |
| Previous major bleeding event | 201 (3.0) | 14 (4.7) | 187 (3.0) | 0.09 |
| **Drug** |  |  |  |  |
| Vitamin K antagonist | 3292 (49.6) | 178 (59.7) | 3114 (49.2) | 0.0004 |
| Direct oral anticoagulant | 3341 (50.4) | 120 (40.3) | 3221 (50.8) | 0.0004 |
| Antiplatelet drug | 1314 (19.8) | 80 (26.9) | 1234 (19.5) | 0.002 |
| **Blood test** |  |  |  |  |
| Hemoglobin, g/dL | 13.7±1.7 | 13.1±1.8 | 13.7±1.7 | <0.0001 |
| Platelet count, ×10<sup>4</sup>/μL | 19.6±5.9 | 19.1±6.5 | 19.6±5.8 | 0.14 |
| Serum creatinine, mg/dL | 0.86 [0.73, 1.02] | 0.89 [0.73, 1.08] | 0.86 [0.73, 1.02] | 0.049 |
| Urinary acid, mg/dL | 5.9±1.5 | 6.0±1.6 | 5.9±1.5 | 0.08 |
| Low-density lipoprotein-cholesterol, mg/dL | 103.6±28.2 | 98.4±28.4 | 103.8±28.1 | 0.001 |
| High-density lipoprotein-cholesterol, mg/dL | 55.0±15.4 | 53.1±15.1 | 55.1±15.4 | 0.03 |
| Triglycerides, mg/dL | 111 [79, 160] | 108 [76, 163] | 112 [80, 160] | 0.17 |
| Hemoglobin A1c, % | 6.0±0.8 | 6.1±0.9 | 6.0±0.8 | 0.48 |
| Brain natriuretic peptide, pg/mL | 107.5 [56.3, 194.7] | 121.2 [63.5, 218.8] | 107.0 [56.2, 193.5] | 0.09 |

### Predictors of major bleeding events

**Table 2** lists the results of univariate and multivariate logistic regression analysis. Age, use of VKA, use of an antiplatelet drug, and serum creatinine were selected by univariate logistic regression analysis (p < 0.05) and entered into the multivariate model. Multivariate regression analysis revealed older age (odds ratio, 1.03; 95% CI, 1.02–1.05; p < 0.0001), VKA (odds ratio, 1.42; 95% CI, 1.11–1.80; p = 0.005), and antiplatelet drug use (odds ratio, 1.31; 95% CI, 1.003– 1.718; p = 0.048) as factors significantly associated with incidence of major bleeding.

**Table 2.** Logistic regression analysis for major bleeding events

|  | Univariable |  |  | Multivariable |  |  |
| --- | --- | --- | --- | --- | --- | --- |
|  | OR | 95% CI | p value | OR | 95% CI | p value |
| Age | 1.04 | 1.02–1.05 | <0.0001 | 1.03 | 1.02–1.05 | <0.0001 |
| Male | 0.99 | 0.77–1.28 | 0.96 |  |  |  |
| Body mass index | 0.97 | 0.08–1.04 | 0.06 |  |  |  |
| Previous major bleeding event | 1.62 | 0.93–2.83 | 0.09 |  |  |  |
| Vitamin K antagonist use | 1.53 | 1.21–1.94 | 0.0004 | 1.42 | 1.11–1.80 | 0.005 |
| Antiplatelet drug use | 1.52 | 1.17–1.97 | 0.002 | 1.31 | 1.003–1.718 | 0.048 |
| Hypertension | 1.23 | 0.94–1.62 | 0.13 |  |  |  |
| Diabetes mellitus | 1.26 | 0.98–1.61 | 0.07 |  |  |  |
| Serum creatinine | 1.18 | 1.04–1.34 | 0.03 | 1.13 | 0.98–1.30 | 0.13 |

95% CI, 95% confidence interval; OR, odds ratio.

### Clinical outcomes

**Table 3** shows the incidences of primary and secondary outcomes. The rate of incidence for each clinical event was significantly higher in patients with major bleeding than in those without. MACCE occurred in 72 patients after major bleeding and in 665 patients without major bleeding (18.33 and 3.22 per 100 patient-years, respectively). The median duration of MACCE after major bleeding was 41 (IQR, 3–300) days. All-cause death occurred in 65 patients after major bleeding and in 464 patients without major bleeding (16.32 and 2.21 per 100 patient-years, respectively). Cardiovascular death occurred in 24 patients after major bleeding and in 192 patients without major bleeding (6.02 and 0.91 per 100 patient-years, respectively). Median duration until death and until cardiovascular death after major bleeding were 43 and 13 days, respectively. Kaplan–Meier analysis showed that occurrences of MACCE and all-cause death were significantly and clearly higher in patients after major bleeding compared with those without major bleeding (both log-rank p <0.0001; **Figures 2, 3**).

**Table 3.**
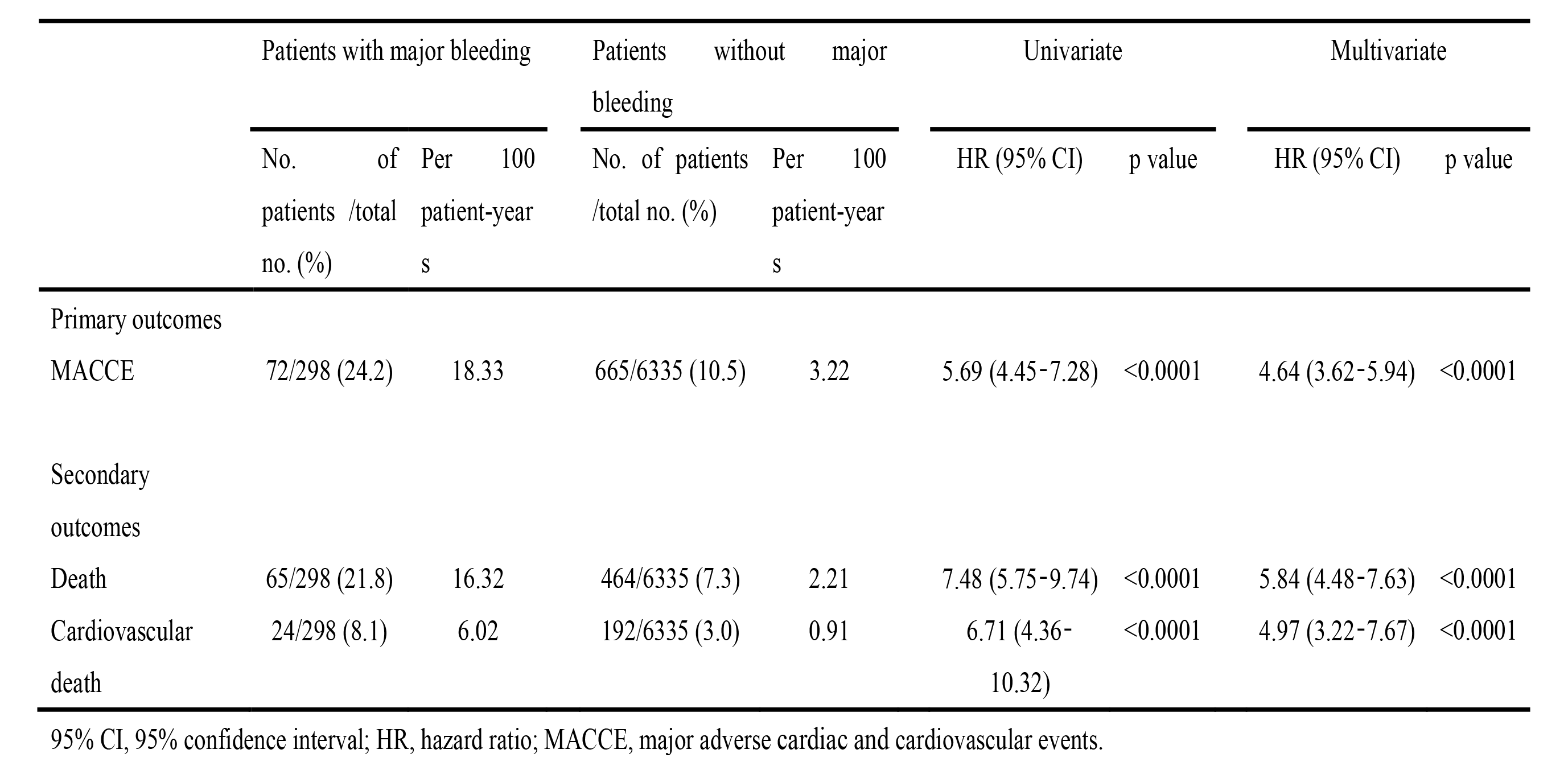
Clinical events and risk after major bleeding events

**Figure 2.**
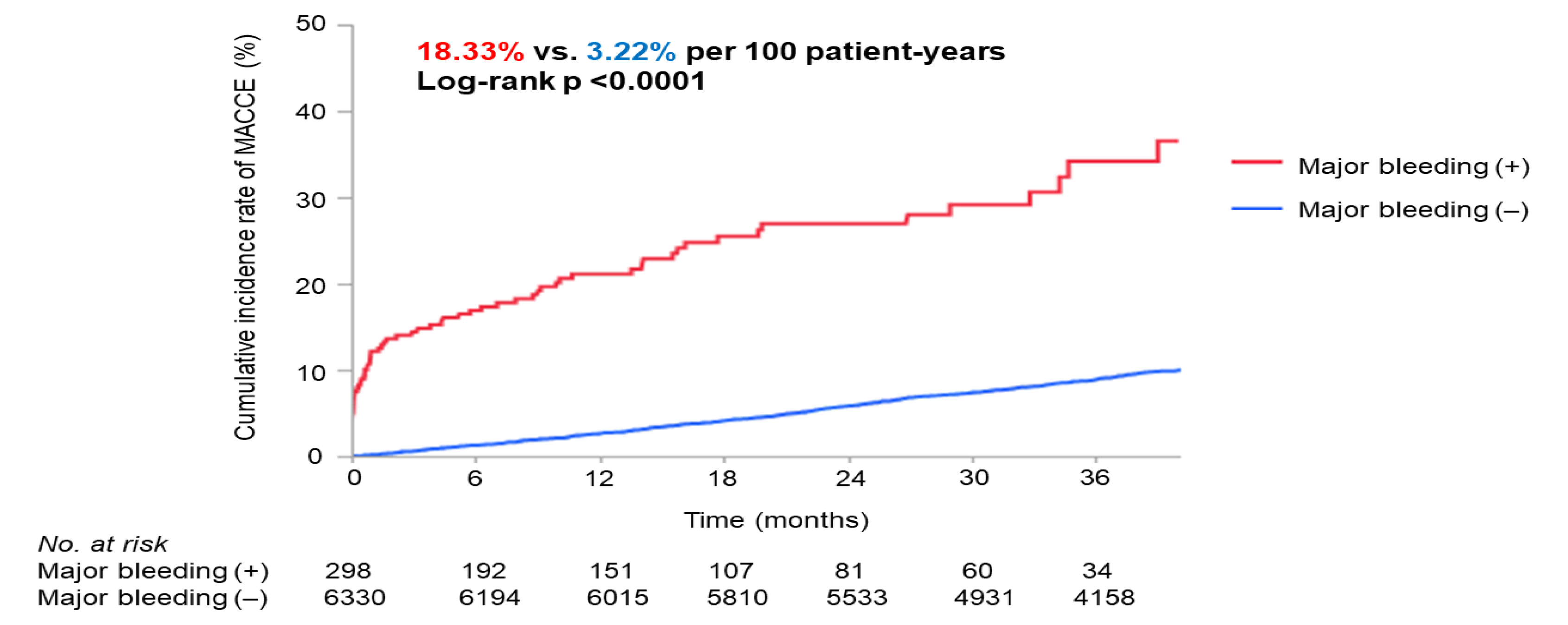
Rates of major adverse cardiac and cerebrovascular events (MACCE) in patients with and without major bleeding Development of MACCE was significantly and clearly higher after major bleeding than without major bleeding (log-rank p <0.0001). MACCE; major adverse cardiac and cerebrovascular events.

**Figure 3.**
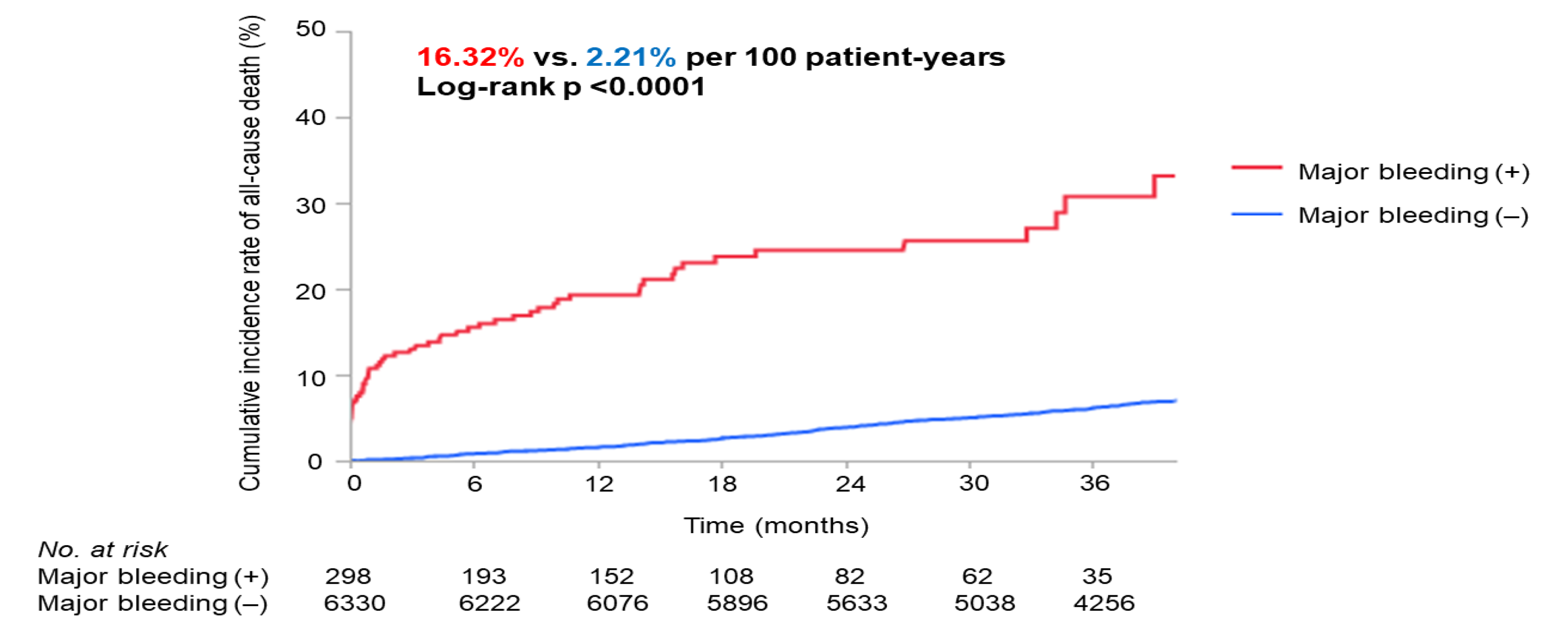
All-cause mortality in patients with and without major bleeding Development of MACCE was significantly and clearly higher after major bleeding than without major bleeding (log-rank p <0.0001)

Multivariate Cox hazard regression analysis showed that risk of MACCE was significantly higher in patients after major bleeding compared with those without major bleeding (HR, 4.64; 95% CI, 3.62–5.94; p <0.0001; **Table 3**). Multivariate Cox hazard analysis showed that major bleeding was also associated with the incidence of all-cause death and of cardiovascular death (HR, 5.84; 95% CI, 4.48–7.63; p <0.0001 and HR, 4.97; 95% CI, 3.22–7.67; p <0.0001, respectively; **Table 3**).

## Discussion

In the present study, we investigated the association between major bleeding and long-term cardiovascular events in patients with AF who were taking OAC. The major findings were as follows: (1) during follow-up, 4.5% of patients (1.38 per 100 patient-years) experienced major bleeding events; (2) older age, VKA use, and antiplatelet drug use were independently associated with incidence of major bleeding events; (3) 24% of patients with major bleeding had MACCE during long-term follow-up (18.33 per 100 patient-years); (4) patients who experienced major bleeding had significantly higher risks of MACCE, all-cause death, and cardiovascular death, and these adverse events occurred relatively early after major bleeding occurred.

As major bleeding events were found to lead to worse clinical outcomes, it is important to pay careful attention to the risk of major bleeding in patients with AF who are taking OACs. A previous report from the ARISTOTLE trial showed that older age, prior bleed, prior stroke, diabetes, lower creatine clearance, decreased hematocrit level, and use of aspirin and nonsteroidal inflammatory drugs were independent predictors of major bleeding among patients with AF ^12^. In addition, recent studies have shown that bleeding risk is higher with VKAs than with DOACs ^13, 14^. Switching to a DOAC should be considered for patients receiving a VKA, particularly in those with high bleeding risk ^13^. The present study found that as well as VKAs, antiplatelet drug use was related to major bleeding events. AF patients with coronary artery disease (CAD) usually take antiplatelet drugs, and 20% of the present patients used these drugs. The AFIRE trial demonstrated that DOAC monotherapy was noninferior to combination therapy with DOAC plus a single antiplatelet agent in terms of efficacy, and was superior in terms of safety in patients with AF and stable CAD ^15^. Therefore, for patients using both OAC and antiplatelet drugs, it is important to evaluate the necessity for continuation of antiplatelet drugs.

Previous reports have shown that major bleeding increased the risk of cardiovascular events in AF patients in the first 30 days after a bleeding event ^7, 12^. Recently, Meyer et al evaluated the long-term risk of adverse clinical outcomes in AF patients with a newly documented bleeding episode ^16^. They reported median time to an adverse outcome event of 142 days after a major bleed. In contrast, the median time until development of MACCE after a major bleed in the present study was only 41 days. Kaikita et al. investigated the association between bleeding and cardiovascular events in AF patients with stable CAD. Their time-adjusted multivariate analysis showed a temporal association between major bleeding and subsequent MACCE, with a particularly high risk of developing MACCE within 1 month after a major bleed ^17^. Even in studies with long-term observation, post-bleeding events occurred relatively early; therefore, careful observation of patients with AF is required during that period.

There are several possible explanations for the increased risk of MACCE after major bleeding. First, bleeding itself is a direct cause of cardiovascular events. Bleeding can be fatal and anemia due to bleeding might lower the threshold for ischemia or heart failure. Acute anemia results in decreased blood supply and sudden hypoxia to the heart. In addition, it exacerbates the preexisting compromised coronary blood supply in patients with CAD, and there is a disproportionate oxygen supply and demand ratio to the heart ^18, 19^. Second, OACs are often discontinued in an effort to control major bleeding ^20^. A previous study has reported that discontinuation of OAC after bleeding episodes was associated with a higher risk of adverse outcomes ^16^. Thus, early resumption of OAC is preferable after hemostasis is achieved and the clinical condition is stabilized, especially in patients with high thrombotic risk. Conversely, restarting OAC after a bleeding event has also been associated with increased risk of recurrent bleeds ^21, 22^. Determining the optimal timing for restarting anticoagulation has the dual therapeutic aims of preventing thrombotic events while minimizing rebleeding, and should be discussed by the medical team. In patients with recurrent severe bleeding and in those who are not able to restart anticoagulation, nonpharmacological therapies such as left atrial appendage closure or occlusion should be considered. Third, the present patients with major bleeding had higher prevalences of ischemic heart disease and prior stroke, and higher CHADS_2_ and HAS-BLED scores than those without major bleeding. In other words, patients at high risk of bleeding not only have an increased risk of bleeding but also an elevated risk of cardiovascular events. Therefore, we should be aware of bleeding as well as cardiovascular events in these patients.

This study had several limitations. First, as a prospective observational study, unknown confounding factors or selection bias might have affected the outcomes, regardless of the analytical adjustments. Second, it was unknown whether patients who had experienced major bleeding events discontinued OACs, and when they restarted anticoagulation therapy. It is also possible that some patients might have changed the type of OAC taken or lowered their dosage to reduce bleeding risks during the course of the study.

## Conclusions

In the present analysis of data from two large-scale prospective registries, major bleeding was associated with long-term adverse cardiovascular events among AF patients taking OACs. Therefore, to improve clinical outcomes in patients with AF, it is important to reduce bleeding risks. When a major bleeding event occurs, patients should be monitored carefully for cardiovascular events, especially in the early phase after bleeding.

## Funding

This work was supported by scholarship funds received from the following (in alphabetical order): Abbott Japan, Astellas Pharma, AstraZeneca, Bayer Healthcare, Boehringer-Ingelheim, Boston Scientific Japan, Bristol-Meyers Squibb, Crosswill Medical, Daiichi-Sankyo, Eisai, Fukuda Denshi, FUJIFILM RI Pharma, Japan Lifeline, Kowa Pharmaceutical, Kyowa Hakko Kirin, Mitsubishi Tanabe Pharma, Medical Hearts, Medtronic Japan, Mochida, MSD, Nippon Shinyaku, Otsuka Pharmaceutical, Pfizer, Philips Respironics, Roche Diagnostics, Sanwa Kagaku Kenkyusho, Sanofi, Shionogi, Sumitomo Dainippon Pharma, Takeda Pharmaceutical, and Nihon Medi-Physics. This study was conducted as investigator-initiated research based on a contract with, and financially supported by, Bayer Yakuhin Ltd.

The funding sources had no roles in the study design; in the collection, analysis, and interpretation of data; in the writing of the report; or in the decision to submit the article for publication.

## Disclosures

Dr. Daida has accepted remuneration from Daiichi-Sankyo, Kowa, MSD, Novartis Pharma, and Bayer Yakuhin, Sanofi K.K., Taisho Pharmaceutical, Abott Medical, Otsuka, Amgen K.K., Pfizer, Fukuda Denshi, Tsumura, Toa Eiyo; received research grants from Philips Japan, Toho Holdings, Asahi Kasei, Fujifilm holdings; Inter Reha, Glory, BMS, Abott, and Boehringer Ingelheim and received scholarship funds from Daiichi-Sankyo, Bayer Yakuhin, and Eisai.

Dr. Matsumoto has lecture’s fee from Nihon Medi-physics and PDRadio Pharma, and has received scholarship donation from PDRadio Pharma.

Dr. Miyauchi has accepted remuneration from Astellas, MSD, Bayer Yakuhin, Sanofi, Daiichi-Sankyo, Boehringer-Ingelheim, and Bristol-Myers Squibb.

Dr. Minamino serves as a consultant to Fukuda Denshi; has accepted remuneration from Kowa, Mitsubishi Tanabe, Daiichi-Sankyo, Bayer Yakuhin, Boehringer-Ingelheim, AstraZeneca KK, Medtronic Japan, Otsuka, Novartis Pharma, Takeda, MSD, Actelion, and Biotronik Japan; accepted research grants from Boehringer-Ingelheim, Bourbon Corporation, and Astellas Pharma; and received scholarship funds from Medical Hearts, Abbott Medical Japan, Japan Lifeline, Medtronic Japan, Mochida, Roche Diagnostics, Takeda, Crosswill Medical, Daiichi-Sankyo, Boehringer-Ingelheim, Boston Scientific Japan, Kowa, Otsuka, Bristol-Myers Squibb, Fukuda Denshi, and Astellas.

Dr. Okumura has received research funding from Bayer Healthcare and Biosense Webster, Inc., scholarship donation from Boston Scientific Japan, and Endowed Courses from Boston Scientific Japan, Japan life line, Fukuda Denshi, Abbott Japan, BIOTRONIK Japan, Medtronic Japan, and has accepted remuneration from Bayer Healthcare, Daiichi-Sankyo, Bristol-Meyers Squibb, AstraZeneca K.K., Ono Pharmaceutical, and Medtronic Japan.

## Data Availability

All data referenced in the manuscript is available.

## Acknowledgements

We wish to express our sincere appreciation to all physicians, coordinators, and patients who participated in this study.

## Notes

### Competing Interest Statement

The authors have declared no competing interest.

### Author Declarations

approval numbers: M11-0799 [11 November 2014] for RAFFINE RK-130111-2 [1 February 2013] for SAKURA

## Reference

1. Benjamin EJ, Muntner P, Alonso A, Bittencourt MS, Callaway CW, Carson AP, Chamberlain AM, Chang AR, Cheng S, Das SR, Delling FN, Djousse L, Elkind MSV, Ferguson JF, Fornage M, Jordan LC, Khan SS, Kissela BM, Knutson KL, Kwan TW, Lackland DT, Lewis TT, Lichtman JH, Longenecker CT, Loop MS, Lutsey PL, Martin SS, Matsushita K, Moran AE, Mussolino ME, O’Flaherty M, Pandey A, Perak AM, Rosamond WD, Roth GA, Sampson UKA, Satou GM, Schroeder EB, Shah SH, Spartano NL, Stokes A, Tirschwell DL, Tsao CW, Turakhia MP, VanWagner LB, Wilkins JT, Wong SS, Virani SS. Heart disease and stroke statistics-2019 update: A report from the american heart association. Circulation. 2019;139:e56–e528

2. Hindricks G, Potpara T, Dagres N, Arbelo E, Bax JJ, Blomström-Lundqvist C, Boriani G, Castella M, Dan GA, Dilaveris PE, Fauchier L, Filippatos G, Kalman JM, La Meir M, Lane DA, Lebeau JP, Lettino M, Lip GYH, Pinto FJ, Thomas GN, Valgimigli M, Van Gelder IC, Van Putte BP, Watkins CL. 2020 esc guidelines for the diagnosis and management of atrial fibrillation developed in collaboration with the european association for cardio-thoracic surgery (eacts): The task force for the diagnosis and management of atrial fibrillation of the european society of cardiology (esc) developed with the special contribution of the european heart rhythm association (ehra) of the esc. European heart journal. 2021;42:373–498

3. Ruff CT, Giugliano RP, Braunwald E, Hoffman EB, Deenadayalu N, Ezekowitz MD, Camm AJ, Weitz JI, Lewis BS, Parkhomenko A, Yamashita T, Antman EM. Comparison of the efficacy and safety of new oral anticoagulants with warfarin in patients with atrial fibrillation: A meta-analysis of randomised trials. *Lancet (London*, England). 2014;383:955–962

4. Connolly SJ, Ezekowitz MD, Yusuf S, Eikelboom J, Oldgren J, Parekh A, Pogue J, Reilly PA, Themeles E, Varrone J, Wang S, Alings M, Xavier D, Zhu J, Diaz R, Lewis BS, Darius H, Diener HC, Joyner CD, Wallentin L. Dabigatran versus warfarin in patients with atrial fibrillation. N Engl J Med. 2009;361:1139–1151

5. Granger CB, Alexander JH, McMurray JJ, Lopes RD, Hylek EM, Hanna M, Al-Khalidi HR, Ansell J, Atar D, Avezum A, Bahit MC, Diaz R, Easton JD, Ezekowitz JA, Flaker G, Garcia D, Geraldes M, Gersh BJ, Golitsyn S, Goto S, Hermosillo AG, Hohnloser SH, Horowitz J, Mohan P, Jansky P, Lewis BS, Lopez-Sendon JL, Pais P, Parkhomenko A, Verheugt FW, Zhu J, Wallentin L. Apixaban versus warfarin in patients with atrial fibrillation. N Engl J Med. 2011;365:981–992

6. Patel MR, Mahaffey KW, Garg J, Pan G, Singer DE, Hacke W, Breithardt G, Halperin JL, Hankey GJ, Piccini JP, Becker RC, Nessel CC, Paolini JF, Berkowitz SD, Fox KA, Califf RM. Rivaroxaban versus warfarin in nonvalvular atrial fibrillation. N Engl J Med. 2011;365:883–891

7. Held C, Hylek EM, Alexander JH, Hanna M, Lopes RD, Wojdyla DM, Thomas L, Al-Khalidi H, Alings M, Xavier D, Ansell J, Goto S, Ruzyllo W, Rosenqvist M, Verheugt FW, Zhu J, Granger CB, Wallentin L. Clinical outcomes and management associated with major bleeding in patients with atrial fibrillation treated with apixaban or warfarin: Insights from the aristotle trial. European heart journal. 2015;36:1264–1272

8. Miyazaki S, Miyauchi K, Hayashi H, Tanaka R, Nojiri S, Miyazaki T, Sumiyoshi M, Suwa S, Nakazato Y, Urabe T, Hattori N, Daida H. Registry of japanese patients with atrial fibrillation focused on anticoagulant therapy in the new era: The raffine registry study design and baseline characteristics. J Cardiol. 2018;71:590–596

9. Okumura Y, Yokoyama K, Matsumoto N, Tachibana E, Kuronuma K, Oiwa K, Matsumoto M, Kojima T, Hanada S, Nomoto K, Arima K, Takahashi F, Kotani T, Ikeya Y, Fukushima S, Itoh S, Kondo K, Chiku M, Ohno Y, Onikura M, Hirayama A, The Sakura Af Registry I. Current use of direct oral anticoagulants for atrial fibrillation in japan: Findings from the sakura af registry. J Arrhythm. 2017;33:289–296

10. Miyazaki S, Miyauchi K, Hayashi H, Yamashiro K, Tanaka R, Nishizaki Y, Nojiri S, Suwa S, Sumiyoshi M, Nakazato Y, Urabe T, Hattori N, Minamino T, Daida H. Trends of anticoagulant use and outcomes of patients with non-valvular atrial fibrillation: Findings from the raffine registry. J Cardiol. 2022;80:41–48

11. Schulman S, Kearon C. Definition of major bleeding in clinical investigations of antihemostatic medicinal products in non-surgical patients. Journal of thrombosis and haemostasis: JTH. 2005;3:692–694

12. Hylek EM, Held C, Alexander JH, Lopes RD, De Caterina R, Wojdyla DM, Huber K, Jansky P, Steg PG, Hanna M, Thomas L, Wallentin L, Granger CB. Major bleeding in patients with atrial fibrillation receiving apixaban or warfarin: The aristotle trial (apixaban for reduction in stroke and other thromboembolic events in atrial fibrillation): Predictors, characteristics, and clinical outcomes. Journal of the American College of Cardiology. 2014;63:2141–2147

13. Carnicelli AP, Hong H, Connolly SJ, Eikelboom J, Giugliano RP, Morrow DA, Patel MR, Wallentin L, Alexander JH, Cecilia Bahit M, Benz AP, Bohula EA, Chao TF, Dyal L, Ezekowitz M, K AAF, Gencer B, Halperin JL, Hijazi Z, Hohnloser SH, Hua K, Hylek E, Toda Kato E, Kuder J, Lopes RD, Mahaffey KW, Oldgren J, Piccini JP, Ruff CT, Steffel J, Wojdyla D, Granger CB. Direct oral anticoagulants versus warfarin in patients with atrial fibrillation: Patient-level network meta-analyses of randomized clinical trials with interaction testing by age and sex. Circulation. 2022;145:242–255

14. Jackevicius CA, Lu L, Ghaznavi Z, Warner AL. Bleeding risk of direct oral anticoagulants in patients with heart failure and atrial fibrillation. Circulation. Cardiovascular quality and outcomes. 2021;14:e007230

15. Yasuda S, Kaikita K, Akao M, Ako J, Matoba T, Nakamura M, Miyauchi K, Hagiwara N, Kimura K, Hirayama A, Matsui K, Ogawa H, Investigators A. Antithrombotic therapy for atrial fibrillation with stable coronary disease. N Engl J Med. 2019;381:1103–1113

16. Meyre PB, Blum S, Hennings E, Aeschbacher S, Reichlin T, Rodondi N, Beer JH, Stauber A, Müller A, Sinnecker T, Moutzouri E, Paladini RE, Moschovitis G, Conte G, Auricchio A, Ramadani A, Schwenkglenks M, Bonati LH, Kühne M, Osswald S, Conen D. Bleeding and ischaemic events after first bleed in anticoagulated atrial fibrillation patients: Risk and timing. European heart journal. 2022;43:4899–4908

17. Kaikita K, Yasuda S, Akao M, Ako J, Matoba T, Nakamura M, Miyauchi K, Hagiwara N, Kimura K, Hirayama A, Matsui K, Ogawa H. Bleeding and subsequent cardiovascular events and death in atrial fibrillation with stable coronary artery disease: Insights from the afire trial. Circ Cardiovasc Interv. 2021;14:e010476

18. Padda J, Khalid K, Hitawala G, Batra N, Pokhriyal S, Mohan A, Cooper AC, Jean-Charles G. Acute anemia and myocardial infarction. Cureus. 2021;13:e17096

19. Wang G, Zhao N, Zhong S, Li J. A systematic review on the triggers and clinical features of type 2 myocardial infarction. Clin Cardiol. 2019;42:1019–1027

20. Tomaselli GF, Mahaffey KW, Cuker A, Dobesh PP, Doherty JU, Eikelboom JW, Florido R, Gluckman TJ, Hucker WJ, Mehran R, Messé SR, Perino AC, Rodriguez F, Sarode R, Siegal DM, Wiggins BS. 2020 acc expert consensus decision pathway on management of bleeding in patients on oral anticoagulants: A report of the american college of cardiology solution set oversight committee. Journal of the American College of Cardiology. 2020;76:594–622

21. Tapaskar N, Ham SA, Micic D, Sengupta N. Restarting warfarin vs direct oral anticoagulants after major gastrointestinal bleeding and associated outcomes in atrial fibrillation: A cohort study. Clin Gastroenterol Hepatol. 2022;20:381–389.e389

22. Tapaskar N, Pang A, Werner DA, Sengupta N. Resuming anticoagulation following hospitalization for gastrointestinal bleeding is associated with reduced thromboembolic events and improved mortality: Results from a systematic review and meta-analysis. Dig Dis Sci. 2021;66:554–566

